# Extending Inferences From A Real-world Clinico-Genomic Database Non-Small Cell Lung Cancer Sample To The Underlying Target Population Using Combined Information From Cancer Registrations

**DOI:** 10.1101/2023.06.15.23291372

**Authors:** Darren S. Thomas, Simon Collin, Luis C. Berrocal-Almanza, Heide Stirnadel-Farrant, Yiduo Zhang, Ping Sun

## Abstract

**Purpose:** The study aimed to extend inferences on the Average Treatment Effects (ATEs) from a Flatiron Health Clinico-Genomic Database (CGDB) Non-Small Cell Cancer (NSCLC) sample to the target population represented by SEER cancer registrations. The work also demonstrates the potential for the non-random selection to cause bias through a quantitative framework that compares the marginal and joint distributions of effect modifiers through each non-random selection process.

**Methods:** ATEs for a binary treatment were estimated within the sample (SATE) and extended to the population (PATE) using combined information from SEER and a weighted estimator to standardize the joint distributions of effect modifiers. To understand potential biases through selection, the marginal and joint distributions of effect modifiers were compared through each stepwise process using two referent populations: SEER registrations and a superset of all NSCLC patients in the Flatiron Health network.

**Results:** Within a subset of 1,166 stage III-IV NSCLCs receiving a binary treatment and combined information from 149,056 SEER registrations, the SATE & PATE for differences in survival at month 48 were −3.7 (−8.7, 1.6) & −3.4 (−8.7, 2.8) percentage points. Through each sequential selection, the joint distributions of effect modifiers were not discernibly different among the selected & unselected. Estimates of survival were unbiased by selection.

**Conclusions:** Combined information from cancer registrations can be used to extend inferences from a selected sample to the target population. ATEs within a CGDB were an unbiased estimate of the population because the sequential selection did not differentially select effect modifiers causative of survival.

**Key Points:**

- Combined information from cancer registrations can be used to extend inferences from a selected sample to the target population
- We outline a quantitative framework for determining the potential for non-random selection to cause bias, through comparing the marginal and joint distributions of effect modifiers through each sequential selection process
- Average Treatment Effects within a highly selected genetic cohort were an unbiased estimate of the population because the sequential selection did not differentially select effect modifiers causative of survival

**Plain Language Summary:** In pharmacoepidemiology we oftentimes learn about the effectiveness of treatments within a smaller sample in attempt to understand how they would work in the broader population. When this sampling is non-random — like when we use Real-World Data in the form of Electronic Medical Records or insurance claims — what we learn in this sample may not translate to the broader population. In this study we show how we can publicly available data in the form of cancer registrations to better understand how these treatments work in the population. We show that what we learn about treatments within our highly selected sample that required patients to undergo expensive genetic testing does in fact translate to the population. We also provide a framework showing why this was the case: patients in our selected sample were very similar in their characteristics to the broader population.

## PURPOSE

Rarely is a study sample randomly drawn from the target population. Real-world data are oftentimes a non-random convenience sample of patients receiving healthcare at centres using an Electronic Medical Record system, covered by a defined insurance policy or enrolled in a disease registry ^1,2^. When treatment effect modifiers influence the selection into a sample, the selected sample & unselected target population become unexchangeable in these effect modifiers. In consequence, even in the absence of other confounding and measurement biases, this non-random selection will in expectation bias the Sample Average Treatment Effect (SATE) away from the Population Average Treatment Effect (PATE)^3–6^. This can arise as a function of two causal structures: a type-I selection bias through restricting on a collider or it’s descendants, or a type-II selection bias through restricting on an effect-measure modifier ^4^. We herein focus on the former; wherein selection on a collider opens a non-causal path from treatment to outcome.

Our motivating example is in the study of the Flatiron Health-Foundation Medicine Clinico-Genomic Database (CGDB) Non-Small Cell Lung Cancer (NSCLC) sample ^7^. The database represents the intersection of NSCLC patients treated within the Flatiron Health Research Network who underwent human technology-assisted chart abstraction and whose tumor biopsy was submitted for Next Generation Sequencing (NGS). The differential selection of patients into the CGDB can therefore be caused by the geographic sampling of Flatiron Health clinics across the USA, or in the requirement for patients to meet further eligibility criteria. Though the selection of patients to undergo chart abstraction is random, NGS at its introduction was not widely adopted by clinical guidelines or covered by insurers ^8^, which could bias the selection of certain effect modifier subgroups.

Drawing inspiration from the transportability analyses used to extend causal inferences from randomised-controlled trial subjects to a pragmatic population via standardization of the joint distribution of effect modifiers in the sample to the underlying target population ^9–11^, we i) demonstrate how to extrapolate the SATE to PATE in circumstances where non-random selection is unavoidable by combining information on baseline covariates from cancer registrations; and ii) outline a quantitative framework in which the marginal & joint distributions of effect modifiers are compared through each stepwise selection process. Two referent datasets were used to assess the selection of effect modifiers through each process. We define the referent population as the population preceding a selection process ^4^. The National Cancer Institute’s Surveillance, Epidemiology, and End Results (SEER) dataset includes cancer registrations for participating states covering 34.6% of the US population. In the absence of USA-wide cancer registrations, we use this as the target population to which we want to extend our inferences. The Flatiron Health Machine Learning Extracted (ML-E) database is a supersample of all patients treated within the Flatiron Health network from which the CGDB is a non-random sample. Selection from SEER to ML-E therefore represents the influence of geographic sampling; from ML-E to CGDB due to the requirement for NGS; and from SEER to CGDB due to both processes combined.

## METHODS

### Flatiron Health Foundation Medicine CGDB (2011–2021)

The CGDB is a USA-wide de-identified database that links, via deterministic matching, Flatiron Health Research Network Electronic Medical Records from ∼ 280 cancer clinics with NGS data from Foundation Medicine’s Comprehensive Genomic Profiling database ^7^. Retrospective longitudinal clinical data were curated via technology-enabled abstraction of variables from structured and unstructured data. The sample was obtained from records covering the period 1 Jan 2011–31 Dec 2021, including patients with: a lung cancer diagnosis (ICD-9-CM 162.x or ICD-10-CM C34.x or C39.9); confirmation of NSCLC origin; two visits on or after 1 Jan 2011; and a NGS test on a biopsy with pathologist-confirmed histology that is consistent with the abstracted tumour type taken anytime from 30 days before the date of initial diagnosis onwards. The clinical vignette used for estimation of SATE & PATE was a subset of stage III—IV patients that received one of two PD-1 inhibitors.

### Flatiron Health ML-E Database (2011–2021)

The ML-E database is a de-identified database of all NSCLC patients receiving care within the Flatiron Health network. The ML-E database uses Machine Learning to extract variables from unstructured clinical data, without confirmation by chart abstraction ^12^. The referent population was obtained from records covering the period 1 Jan 2011–31 Dec 2021. NSCLC case inclusion criteria were the same as for the CGDB sample except for the requirement for a NGS.

### SEER Incidence Data (2011-2016)

The SEER program curates demographic and clinical data on cancers registered by participating state-level registries covering 34.6% of the USA population ^13^. A target population was created from publicly available de-identified SEER registrations from the period 1 Jan 2011-31 Dec 2016 by filtering for case listings with histologically confirmed NSCLC defined by ICD-9-CM/ICD-10-CM and relevant ICD-O-2 histology (Supporting Table 1).

### Study Design

The study aimed to transport average treatment effects from the CGDB sample to the target population represented by SEER. The secondary aims were to compare the marginal and joint distributions of effect modifiers through each selection process and to compare how a transportability-weighted estimator influences Real-world Overall Survival (rwOS). Each analysis concatenate two datasets (the sample and upstream referent population) for a composite dataset, with an indicator variable indicating selection into the sample. The set of baseline effect modifiers, *Z*, was: age [at diagnosis], gender, race, stage (AJCC 7/8^th^ edition for CGDB & ML-E or SEER stage for SEER), and histological classification. The SEER stages Regional, Localized, and Distant are broadly equivalent to AJCC stages I—II, III, and IV respectively. Gender was unrecorded for SEER therefore we used biological sex as a proxy. Patients with incomplete data were excluded.

### Identifiability of Causal Estimand

The causal estimand of interest was the Average Treatment Effect at times *t* (*ATE*_*t*_) as a difference in potential outcomes of survival under initiating a binary treatment *A*, regardless of treatment adherence or discontinuation (intention-to-treat analog). The *ATE*_*t*_ therefore equals *E[Y*_*t*_^*a=1*^ *– Y*_*t*_^*a=0*^], where the *E[Y*^*a*^] represents the potential outcome under treatment *A=1* or *A=0*. For a binary outcome of survival, we take the complement of *E[Y*^*a*^] = *Pr[Y*^*a*^ *= 1]*. The *ATE*_*t*_ was calculated within the CGDB sample (*SATE*_*t*_ *= E[Y*_*t*_^*a=1*^ – *Y*_*t*_^=0^| *S = 1]*, where *S* indicates selection into the sample). Estimation of the SATE under non-randomised treatment assignment are identifiable under the conditions of i) consistency of counterfactual outcomes under treatment *A*; ii) conditional exchangeability over treatment *A* within the selected sample; and iii) positivity of treatment *A* in the selected sample ^9,14^. To extend inferences to the target population (*PATE*_*t*_ *= E[Y*_*t*_^*a=1*^ – *Y*_*t*_^a=0^| *S = 0]*) additional assumptions are required ^9^; namely iv) exchangeability between sample & target populations under selection *S*; and v) positivity of selection *S*. Our causal assumptions in the Supporting File 1 depicts two non-causal paths: a confounding path (*A* < *Z* > *Y*) and a type-1 selection bias path (*A* > [*S*] < *Z* > *Y*).

### Sampling Weights

Under non-nested sampling owing to either the absence of population-wide cancer registration or data privacy preventing the identifiability of CGDB patients within the ML-E, extending inferences necessitates transportation through odds weights ^11,15^. Stabilised Odds of Selection Weights (*W*^*S*^) were estimated from a logistic binomial model trained on the outcome indicating selection (1 for sample, 0 for referent population), conditional on a set of baseline variables (*Z*) causative of selection. Age was modelled with a natural cubic spline with 3 knots. *W*^*S*^ were calculated as:

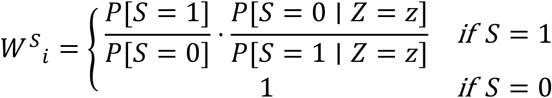

For estimation of the ATEs, Stabilized Probability of Treatment Weights (*W*^*A*^) were calculated within the subset of CGDB patients as:

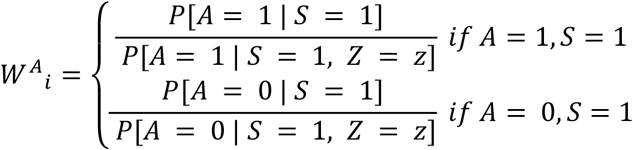

Where both transportability and treatment weights are required for the PATE product weights was used (*W*^*A*^*W*^*S*^).

### Distributions of Effect Modifiers

The marginal distributions of effect modifiers, *Z*, between the selected (unweighted & *S*^*W*^-weighted) and unselected referent populations (ML-E & SEER) were assessed via the Absolute Standardised Difference (ASD), using a threshold of ASD < 0.1 to access balance ^16^.

The joint distributions of effect modifiers were assessed using the Bhattacharyya coefficient (*β*). *β* quantifies the likeness of two probability distributions ^17^; which can be the conditional probabilities under generalizations or conditional odds under transportability. Its value, bounded between 0 and 1 signifying a scale between two disjoint to identical distributions, is proportional to a greater expectation that the SATE will equal the PATE owing to similar distributions of effect modifiers, and in less reliance of extrapolation of the counterfactual under weighting. *β* is calculated using the conditional distribution of selection for the sample (*f*_s_(*x*)) & population (*f*_p_(*x*)) as:

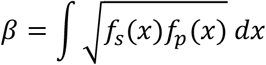

### Real-world Overall Survival

rwOS was calculated for the CGDB sample via Kaplan-Meier estimation. Time zero was the date of diagnosis for analyses of stages I–IV & I–II and the date of initiating first-line systemic therapy for the SATE & PATE and stages III—IV. Entry was delayed until the day of earliest eligibility on receiving NGS results relative to the index date (risk-set adjustment) ^6,18^. The endpoint was a real-world proxy of mortality ^19^. Surviving patients were censored on the latest date of clinical activity. The SATE and PATE were calculated at 12, 24, 36, and 48 months with 95% confidence intervals for the difference in survival calculated from 1,000 bootstraps. For marginal survival, inferences were based on robust variance estimation and a weighted log-rank test was used to compare event times within the CGDB (unweighted vs. *W*^*S*^-weighted), using a pre-specified α of two-sided *P* < 0.00833 (α of 0.05 Bonferroni adjusted for six comparisons).

### Sensitivity Analysis

A sensitivity analysis aligning the study periods of CGDB & SEER (1 Jan 2011–31 Dec 2016) is presented in Supporting Table 11.

### Software

All analyses were undertaken with R version 4.3.1 ^20^.

## RESULTS

### Populations

There were 17,230, 199,278, and 240,943 NSCLC patients in the CGDB, ML-E, and SEER databases (Supporting Table 2). Relative to SEER, the CGDB & ML-E had higher rates of missingness in capturing race (9.0% for CGDB, 9.6% for ML-E, 0.3% for SEER) and stage (3.7% for CGDB, 5.9% for ML-E, 2.5% for SEER), whereas SEER had higher rates of missingness for histological classification (4.0% for CGDB, 4.9% for ML-E, 15.7% for SEER). The respective data within stage I–II & III–IV subsets are presented in Supporting Tables 3 & 4. After exclusion of missing data, there were 14,545, 162,577, and 198,741 remaining in the CGDB, ML-E, and SEER populations.

### Average Treatment Effects

The SATE was estimated within the subset of 1,166 stage III-IV CGDB patients that initiated one of two PD-1 inhibitors using *W*^*a*^ weights, and then extrapolated to the SEER population of 149,056 patients with *W*^*A*^*W*^*S*^ weights. Within the sample there were differences in the ASDs for age, gender, stage, and histology between patients who initiated either PD-1 inhibitor (Table 1). The CGDB sample differed only in race relative to the SEER population (ASD 0.31). The *β* for the joint distributions was 0.97. Relative to the SATEs, the PATEs revised point estimates by + 1.3 (−7.7 vs. −9.0), + 1.0 (−9.8 vs. −10.8), + 1.0 (−3.6 vs. −4.6), and + 0.3 percentage points (−3.4 vs. −3.7) at months 12, 24, 36, and 48, respectively (Table 2). The bootstrap intervals were largely concordant with the exception at month 12 where the PATE suggests a potential change of sign not seen in the SATE. The corresponding counterfactual survival curves are presented in Figure 1.

**Figure 1:**
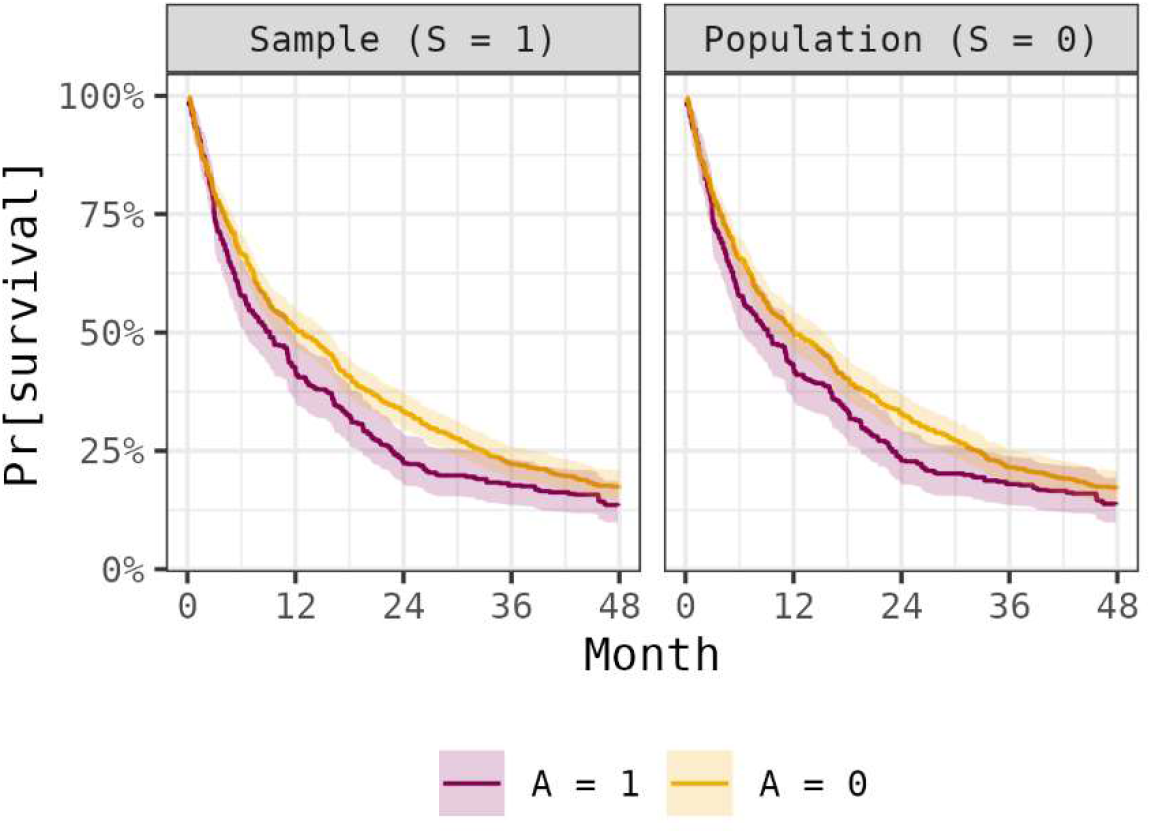
Counterfactual Real-world Overall Survival For Stage III-IV CGDB Patients Receiving a Binary Treatment *A* Estimated Within The Selected Sample And Extrapolated To The SEER Target Population. Curves for the sample were calculated from a *W*^*a*^-weighted estimator and extrapolated to the population with *W*^*A*^*W*^*S*^.

**Table 1:** Effect Modifiers for the Subset of Stage III—IV Patients That Received One of Two PD-1 Inhibitors and an Equally-Bounded SEER Population.

| Characteristic | CGDB Sample<br>(S = 1) |  |  |  | SEER Population<br>(S = 0) |  |
| --- | --- | --- | --- | --- | --- | --- |
|  | All<br>(n 1,166) | A = 1<br>(n 351) | A = 0<br>(n 815) | ASD<sup>1</sup> | All<br>(n 149,056) | ASD<sup>2</sup> |
| Age at diagnosis<sup>3</sup> | 70.0<br>(63.0, 77.0) | 69.0<br>(62.0, 75.0) | 71.0<br>(63.0, 78.0) | 0.19 | 69.0<br>(61.0, 77.0) | 0.03 |
| Gender |  |  |  | 0.12 |  | 0.08 |
| Female | 587<br>(50.3%) | 162<br>(46.2%) | 425<br>(52.1%) |  | 69,327<br>(46.5%) |  |
| Male | 579<br>(49.7%) | 189<br>(53.8%) | 390<br>(47.9%) |  | 79,729<br>(53.5%) |  |
| Race |  |  |  | 0.03 |  | 0.31 |
| Black | 82<br>(7.0%) | 23<br>(6.6%) | 59<br>(7.2%) |  | 18,405<br>(12.3%) |  |
| Other | 197<br>(16.9%) | 60<br>(17.1%) | 137<br>(16.8%) |  | 12,126<br>(8.1%) |  |
| White | 887<br>(76.1%) | 268<br>(76.4%) | 619<br>(76.0%) |  | 118,525<br>(79.5%) |  |
| SEER stage |  |  |  | 0.79 |  | 0.03 |
| Regional | 369<br>(31.6%) | 199<br>(56.7%) | 170<br>(20.9%) |  | 49,014<br>(32.9%) |  |
| Distant | 797<br>(68.4%) | 152<br>(43.3%) | 645<br>(79.1%) |  | 100,042<br>(67.1%) |  |
| Histological classification |  |  |  | 0.21 |  | 0.09 |
| Non-squamous cell carcinoma | 877<br>(75.2%) | 241<br>(68.7%) | 636<br>(78.0%) |  | 105,969<br>(71.1%) |  |
| Squamous cell carcinoma | 289<br>(24.8%) | 110<br>(31.3%) | 179<br>(22.0%) |  | 43,087<br>(28.9%) |  |

Abbreviations: CGDB, Clinico-Genomic Database [population]; SEER, Surveillance, Epidemiology, and End Results [population]; ASD, Absolute Standardized Difference
<sup>1</sup> A = 1 | S = 1 vs. A = 0 | S = 1
<sup>2</sup> S = 1 vs. S = 0
<sup>3</sup> Median (Interquartile range)

**Table 2:**
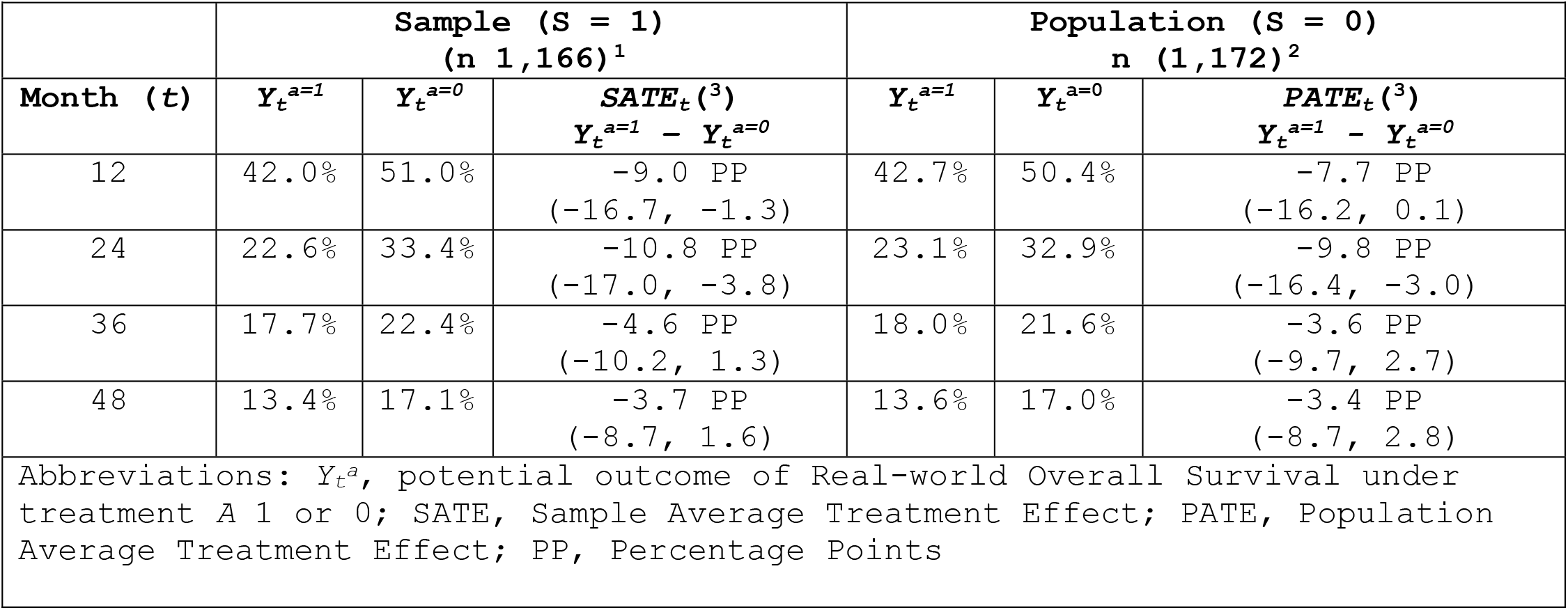
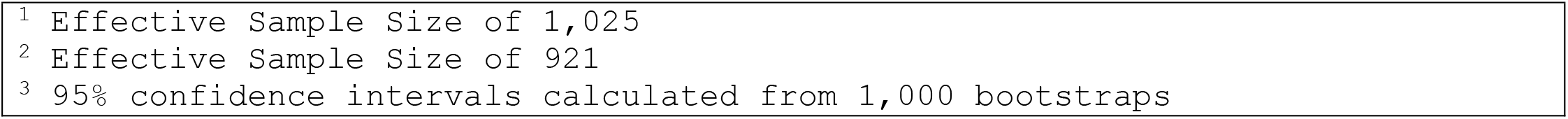
Average Treatment Effects in the Sample and Extended to the Population.

| Month (t) | Sample (S = 1)<br>(n 1,166)<sup>1</sup> |  |  | Population (S = 0)<br>n (1,172)<sup>2</sup> |  |  |
| --- | --- | --- | --- | --- | --- | --- |
| | $Y_t^{a=1}$ | $Y_t^{a=0}$ | $SATE_t^{(3)}$<br>$Y_t^{a=1} - Y_t^{a=0}$ | $Y_t^{a=1}$ | $Y_t^{a=0}$ | $PATE_t^{(3)}$<br>$Y_t^{a=1} - Y_t^{a=0}$ |
| 12 | 42.0% | 51.0% | -9.0 PP<br>(-16.7, -1.3) | 42.7% | 50.4% | -7.7 PP<br>(-16.2, 0.1) |
| 24 | 22.6% | 33.4% | -10.8 PP<br>(-17.0, -3.8) | 23.1% | 32.9% | -9.8 PP<br>(-16.4, -3.0) |
| 36 | 17.7% | 22.4% | -4.6 PP<br>(-10.2, 1.3) | 18.0% | 21.6% | -3.6 PP<br>(-9.7, 2.7) |
| 48 | 13.4% | 17.1% | -3.7 PP<br>(-8.7, 1.6) | 13.6% | 17.0% | -3.4 PP<br>(-8.7, 2.8) |

Abbreviations: $Y_t^a$ , potential outcome of Real-world Overall Survival under treatment A 1 or 0; SATE, Sample Average Treatment Effect; PATE, Population Average Treatment Effect; PP, Percentage Points
<sup>1</sup> Effective Sample Size of 1,025
<sup>2</sup> Effective Sample Size of 921
<sup>3</sup> 95% confidence intervals calculated from 1,000 bootstraps

### Biases Through Selection

The ML-E and SEER populations for stages I–IV, I–II, and III–IV are presented in Table 3 and Supporting Tables 5 & 6. Based on the ASDs (Figure 2), the ML-E population differed from SEER in the distributions of race (ASD 0.28) and stage (ASD 0.21). Subsets defined by stages I–II differed in the composition of race (ASD 0.30) and in stages III-IV by age (ASD 0.17), race (ASD 0.28), and stage (ASD 0.18). *β* for the joint distributions were 0.98, 0.97, and 0.97 for stages I-IV, I–II, and III–IV (Figure 3).

**Table 3:**
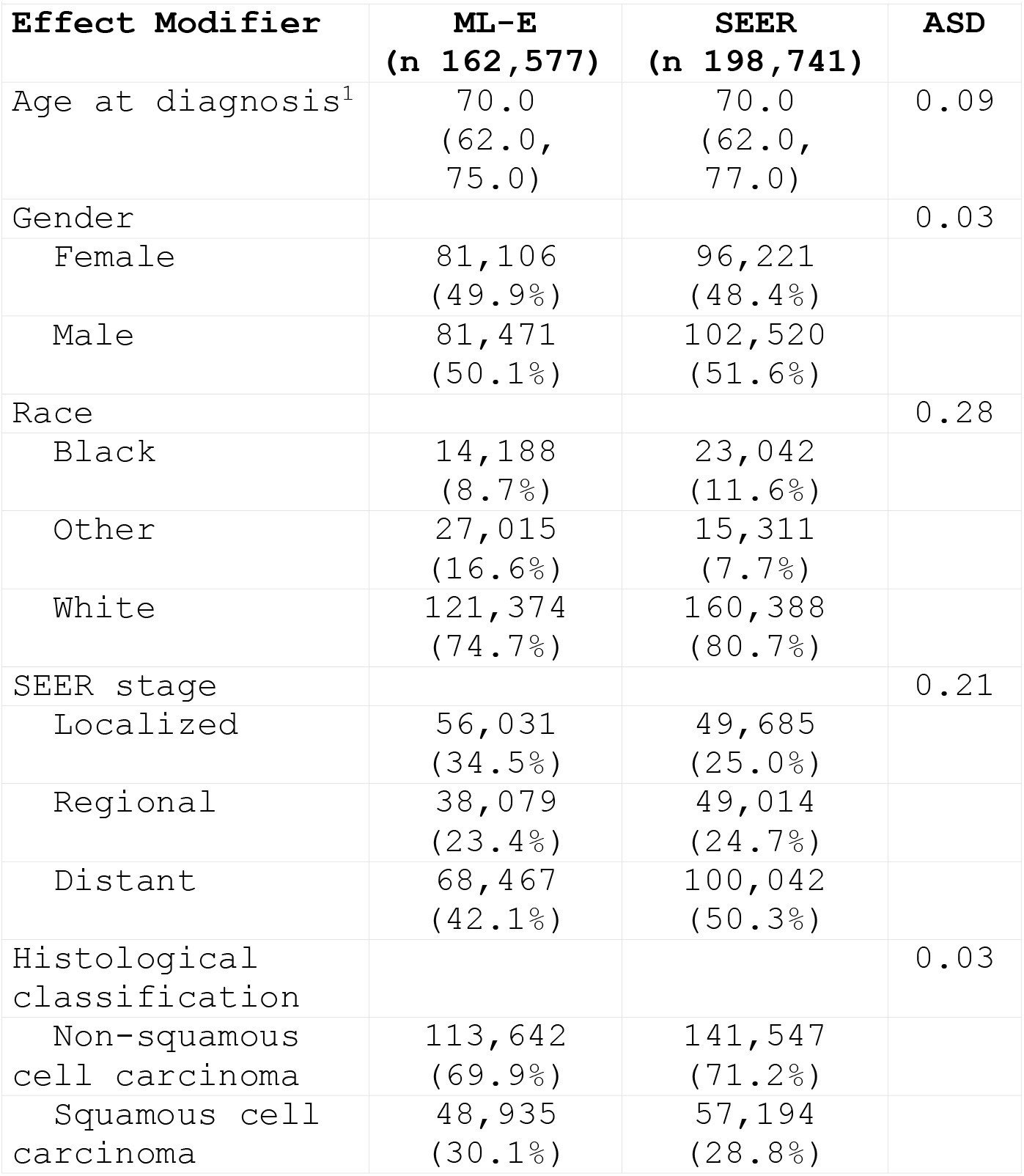
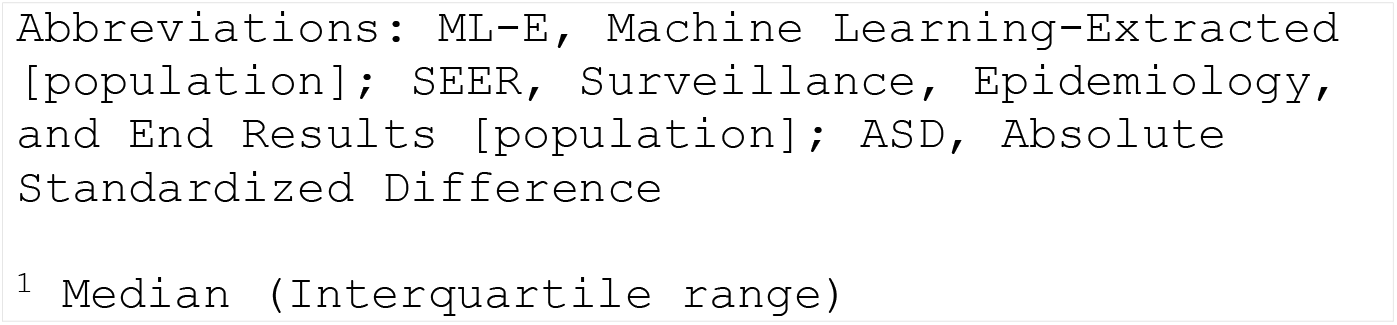
Effect Modifiers for Stages I–IV ML-E & SEER Populations.

| Effect Modifier | ML-E<br>(n 162,577) | SEER<br>(n 198,741) | ASD |
| --- | --- | --- | --- |
| Age at diagnosis<sup>1</sup> | 70.0<br>(62.0,<br>75.0) | 70.0<br>(62.0,<br>77.0) | 0.09 |
| Gender |  |  | 0.03 |
| Female | 81,106<br>(49.9%) | 96,221<br>(48.4%) |  |
| Male | 81,471<br>(50.1%) | 102,520<br>(51.6%) |  |
| Race |  |  | 0.28 |
| Black | 14,188<br>(8.7%) | 23,042<br>(11.6%) |  |
| Other | 27,015<br>(16.6%) | 15,311<br>(7.7%) |  |
| White | 121,374<br>(74.7%) | 160,388<br>(80.7%) |  |
| SEER stage |  |  | 0.21 |
| Localized | 56,031<br>(34.5%) | 49,685<br>(25.0%) |  |
| Regional | 38,079<br>(23.4%) | 49,014<br>(24.7%) |  |
| Distant | 68,467<br>(42.1%) | 100,042<br>(50.3%) |  |
| Histological classification |  |  | 0.03 |
| Non-squamous cell carcinoma | 113,642<br>(69.9%) | 141,547<br>(71.2%) |  |
| Squamous cell carcinoma | 48,935<br>(30.1%) | 57,194<br>(28.8%) |  |

Abbreviations: ML-E, Machine Learning-Extracted [population]; SEER, Surveillance, Epidemiology, and End Results [population]; ASD, Absolute Standardized Difference
<sup>1</sup> Median (Interquartile range)

**Figure 2:**
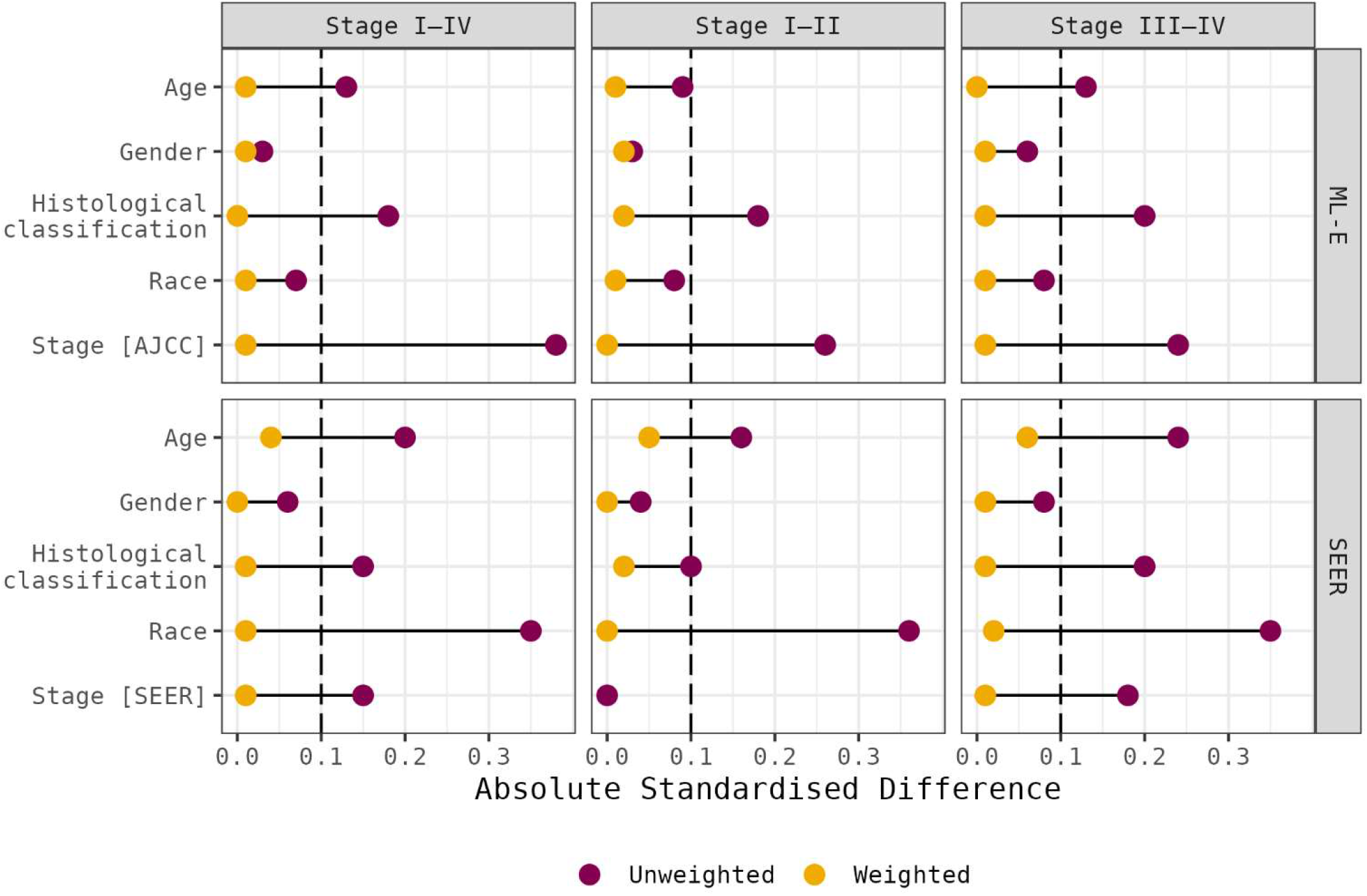
Absolute Standardised Differences Measuring The Differences In The Marginal Distributions of Effect Modifiers Between The Selected CGDB Sample And Unselected ML-E/SEER Referent Populations, Before And After *W*^*S*^ Sampling. An ASD < 0.1 suggests balance in that variable.

**Figure 3:**
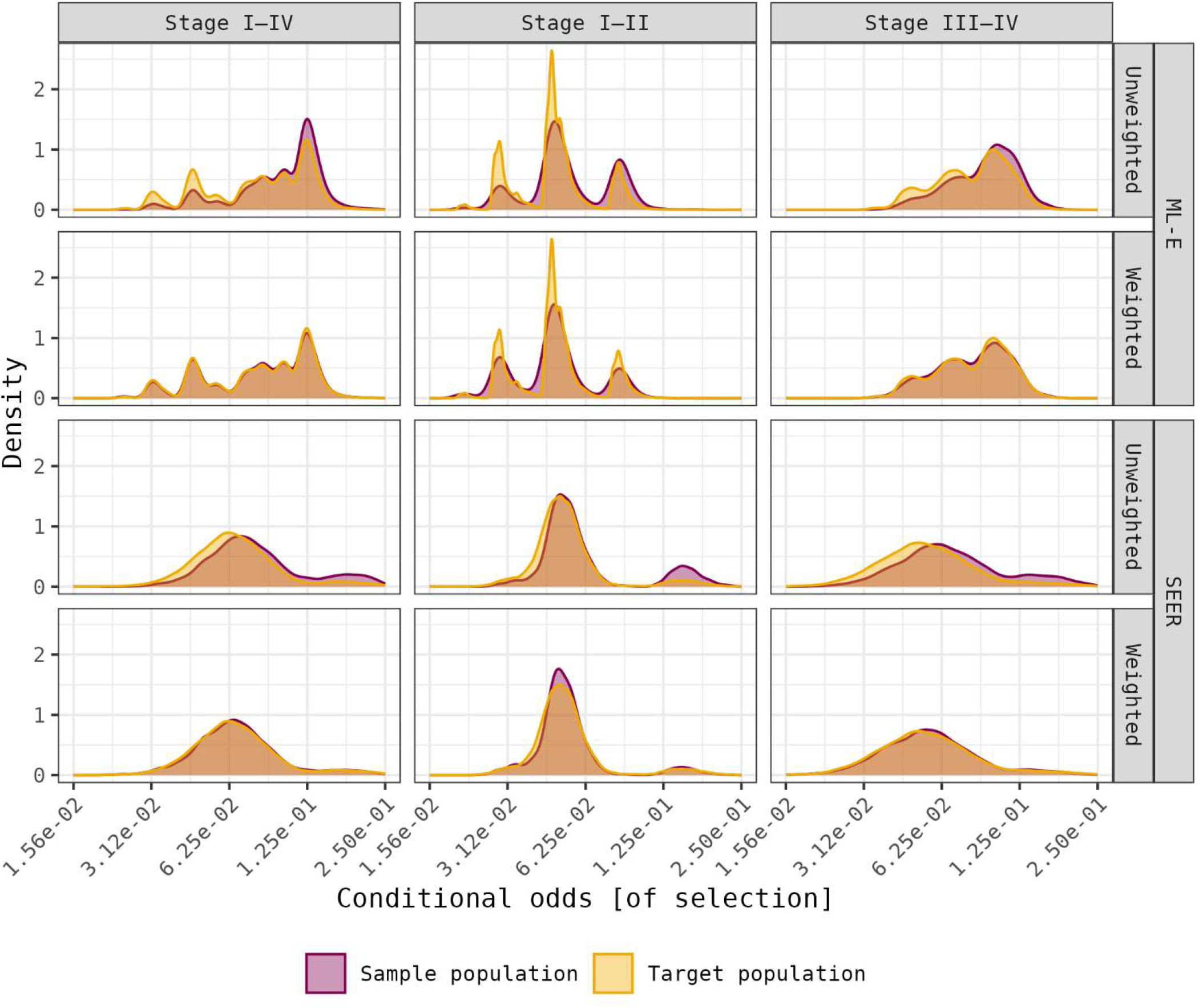
Kernel Densities For The Estimated Odds Of Sample Selection Conditional On A Set Of Effect Modifiers, Before And After *W*^*S*^ Sampling, Using The ML-E Or SEER As The Referent Population.

The CGDB and ML-E populations for stages I–IV, I–II, and III–IV are presented in Table 4 and Supporting Tables 7 & 8 respectively. Based on the ASDs (Figure 2), CGDB patients were younger (ASD 0.13) and had different distributions of AJCC stage (ASD 0.38) and histological classification (ASD 0.18) than all patients who underwent ML-E. Within the subset of stages I–II, CGDB patients differed in the distribution in AJCC stage (ASD 0.26) and histological classification (ASD 0.18). Within the subset of stages III–IV, CGDB patients differed in the distribution in age (ASD 0.13), AJCC stage (ASD 0.24), and histological classification (ASD 0.20). *β* were 0.98, 0.98, and 0.99 for stages I—IV, I—II, and III—IV (Figure 3).

**Table 4:**
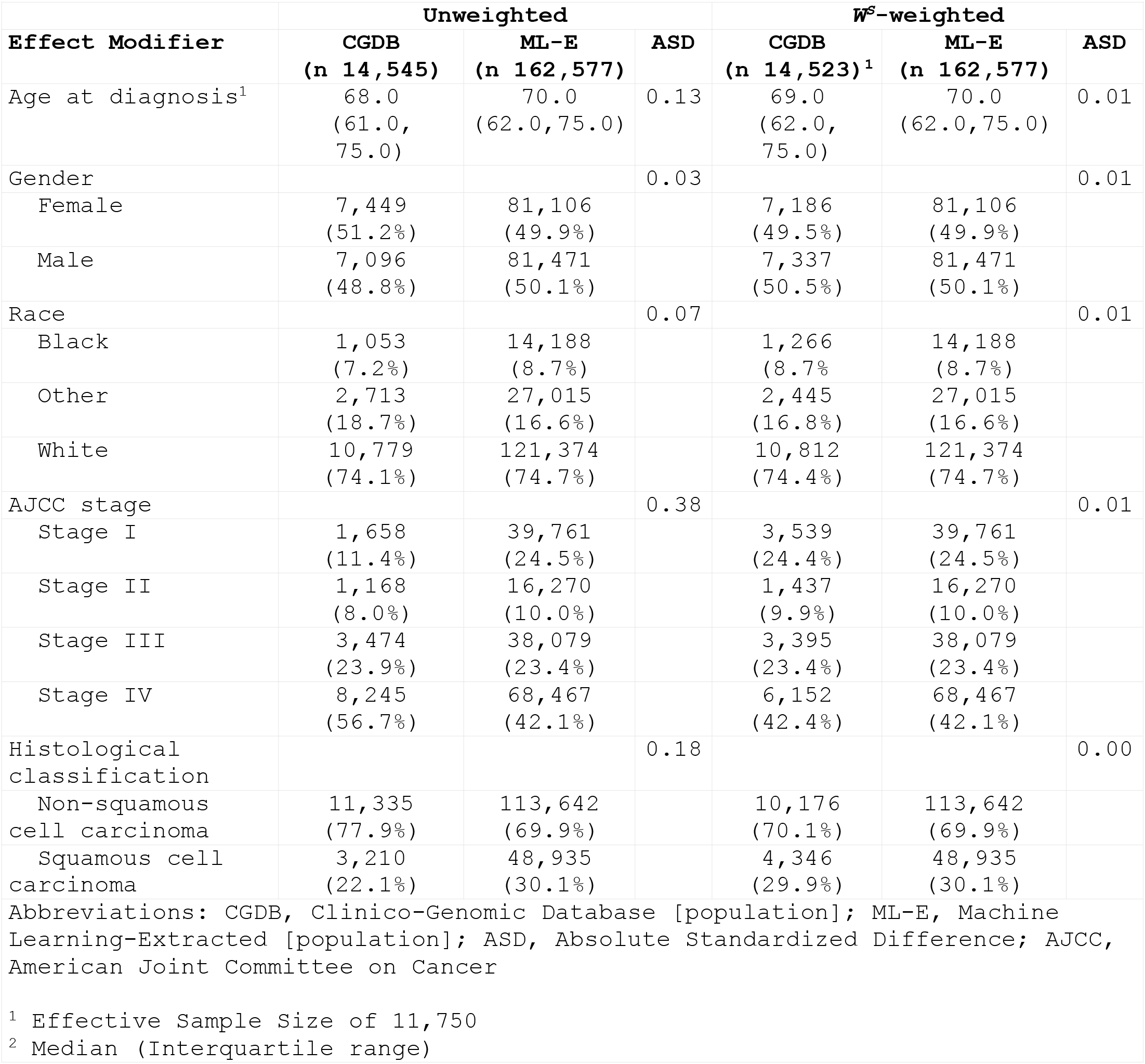
Effect Modifiers for Stages I–IV CGDB & ML-E Populations.

The CGDB and SEER populations for stages I–IV, I–II, and III–IV are presented in Table 5 and Supporting Tables 9 & 10. CGDB patients were younger (ASD 0.20) and had different distributions of race (ASD 0.35), SEER stage (ASD 0.15), and histological classification (ASD 0.15) than SEER registrations. The subset of stages I–II differed in the distributions of age (ASD 0.16), race (ASD 0.36), and histological classification (ASD 0.10), while stages III–IV differed in age (ASD 0.24), race (ASD 0.35), SEER stage (ASD 0.18), and histological classification (ASD 0.20). *β* for stages I—IV, I—II, and III—IV were all 0.97 (Figure 3).

**Table 5:**
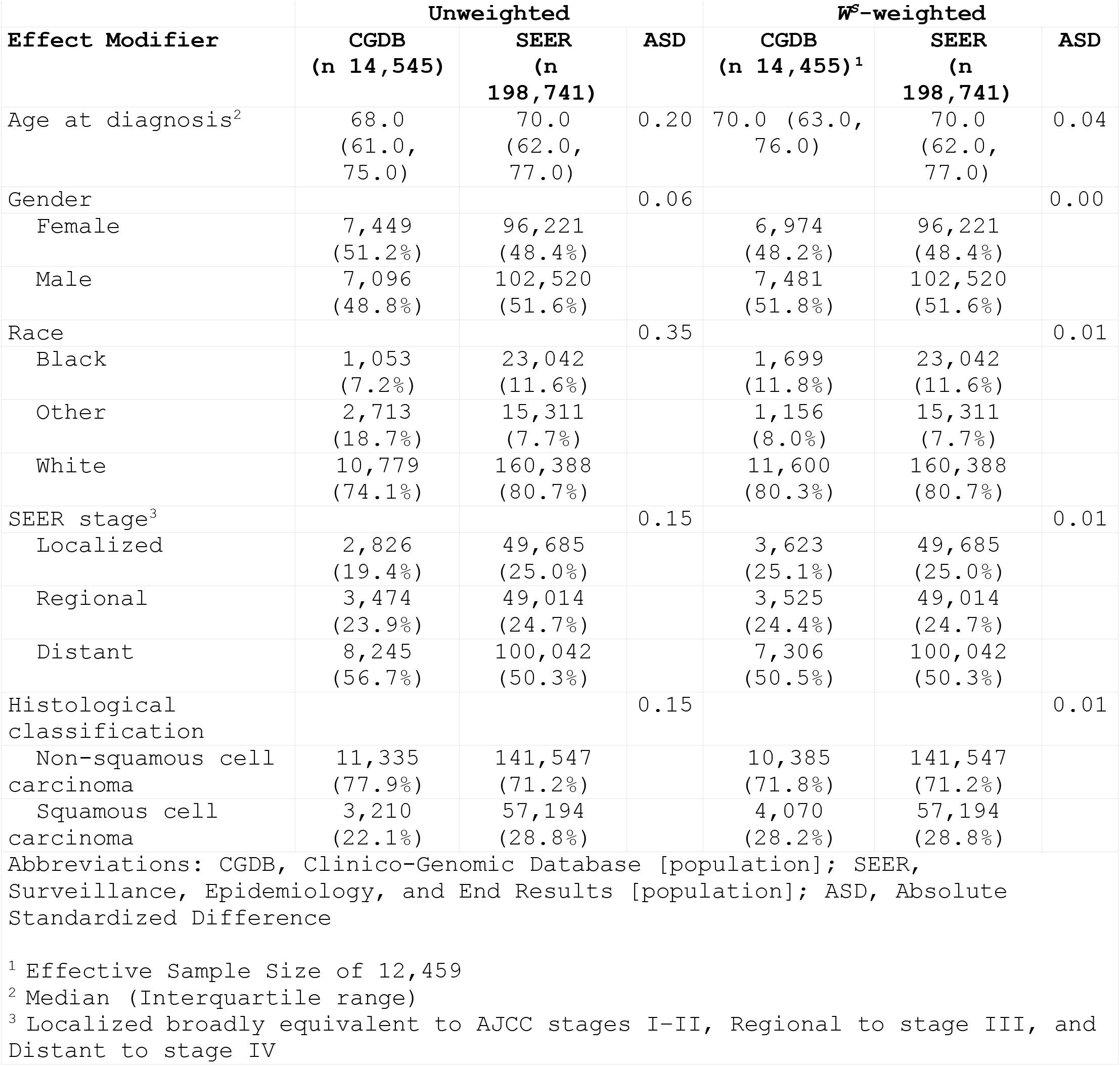
Effect Modifiers for Stages I–IV CGDB & SEER Populations.

| Effect Modifier | Unweighted | | | $W^S$-weighted | | |
| --- | --- | --- | --- | --- | --- | --- |
|  | CGDB<br>(n 14,545) | SEER<br>(n 198,741) | ASD | CGDB<br>(n 14,455)<sup>1</sup> | SEER<br>(n 198,741) | ASD |
| Age at diagnosis<sup>2</sup> | 68.0<br>(61.0,<br>75.0) | 70.0<br>(62.0,<br>77.0) | 0.20 | 70.0 (63.0,<br>76.0) | 70.0<br>(62.0,<br>77.0) | 0.04 |
| Gender |  |  | 0.06 |  |  | 0.00 |
| Female | 7,449<br>(51.2%) | 96,221<br>(48.4%) |  | 6,974<br>(48.2%) | 96,221<br>(48.4%) |  |
| Male | 7,096<br>(48.8%) | 102,520<br>(51.6%) |  | 7,481<br>(51.8%) | 102,520<br>(51.6%) |  |
| Race |  |  | 0.35 |  |  | 0.01 |
| Black | 1,053<br>(7.2%) | 23,042<br>(11.6%) |  | 1,699<br>(11.8%) | 23,042<br>(11.6%) |  |
| Other | 2,713<br>(18.7%) | 15,311<br>(7.7%) |  | 1,156<br>(8.0%) | 15,311<br>(7.7%) |  |
| White | 10,779<br>(74.1%) | 160,388<br>(80.7%) |  | 11,600<br>(80.3%) | 160,388<br>(80.7%) |  |
| SEER stage<sup>3</sup> |  |  | 0.15 |  |  | 0.01 |
| Localized | 2,826<br>(19.4%) | 49,685<br>(25.0%) |  | 3,623<br>(25.1%) | 49,685<br>(25.0%) |  |
| Regional | 3,474<br>(23.9%) | 49,014<br>(24.7%) |  | 3,525<br>(24.4%) | 49,014<br>(24.7%) |  |
| Distant | 8,245<br>(56.7%) | 100,042<br>(50.3%) |  | 7,306<br>(50.5%) | 100,042<br>(50.3%) |  |
| Histological classification |  |  | 0.15 |  |  | 0.01 |
| Non-squamous cell carcinoma | 11,335<br>(77.9%) | 141,547<br>(71.2%) |  | 10,385<br>(71.8%) | 141,547<br>(71.2%) |  |
| Squamous cell carcinoma | 3,210<br>(22.1%) | 57,194<br>(28.8%) |  | 4,070<br>(28.2%) | 57,194<br>(28.8%) |  |

Abbreviations: CGDB, Clinico-Genomic Database [population]; SEER, Surveillance, Epidemiology, and End Results [population]; ASD, Absolute Standardized Difference
<sup>1</sup> Effective Sample Size of 12,459
<sup>2</sup> Median (Interquartile range)
<sup>3</sup> Localized broadly equivalent to AJCC stages I-II, Regional to stage III, and Distant to stage IV

### Influence on Real-world Overall Survival

Sampling with *W*^*S*^ reduced the differences in both variables that were imbalanced (0.1 ≤ ASD) and balanced (ASD < 0.1) (Figure 2). In consequence, *W*^*S*^-weighted samples were balanced with ASDs between 0.01–0.03. The kernel densities for the joint distributions in the unweighted and *W*^*S*^-weighted are presented in Figure 3. Kaplan-Meier estimation of time-conditional rwOS in the unweighted and *W*^*S*^-weighted samples is presented in Figure 4. The corresponding estimates of median survival presented in Table 6 shows that weighting only slightly changed the point estimates and confidence intervals. There were no statistical differences in the distributions of event times for the unweighted and *W*^*S*^-weighted at a prespecified adjusted α of 0.00833.

**Table 6:** Estimates Of Median Real-world Overall Survival And Comparisons Of Event-Time Distributions.

|  | Median Real-world Overall Survival<br>(95% confidence intervals) |  | Event times |
| --- | --- | --- | --- |
| | Unweighted | $W^S$-weighted | Log-rank $P$ value<sup>1</sup> |
| **ML-E as referent population** |  |  |  |
| Stages I-IV<sup>2</sup> | 9.8 (8.9–10.8) | 10.6 (9.7–11.4) | 0.810 |
| Stages I-II<sup>2</sup> | 21.9 (20.5–23.8) | 21.6 (20.1–23.4) | 0.245 |
| Stages III-IV<sup>3</sup> | 11.7 (11.2–12.2) | 11.4 (10.9–11.9) | 0.115 |
| **SEER as referent population** |  |  |  |
| Stages I-IV<sup>2</sup> | 9.8 (8.9–10.8) | 9.5 (8.5–10.5) | 0.035 |
| Stages I-II<sup>2</sup> | 21.9 (20.5–23.8) | 20.7 (19.3–22.4) | 0.158 |
| Stages III-IV<sup>3</sup> | 11.7 (11.2–12.2) | 11.0 (10.6–11.6) | 0.011 |
| Abbreviations: ML-E, Machine Learning-Extracted [population]; SEER, Surveillance, Epidemiology, and End Results [population] |  |  |  |
| <sup>1</sup> prespecified $\alpha$ of 0.00833 (0.05 Bonferroni-adjusted for six comparisons) | | | |
| <sup>2</sup> from date of diagnosis |  |  |  |
| <sup>3</sup> from date of initiating first-line systemic therapy |  |  |  |

**Figure 4:**
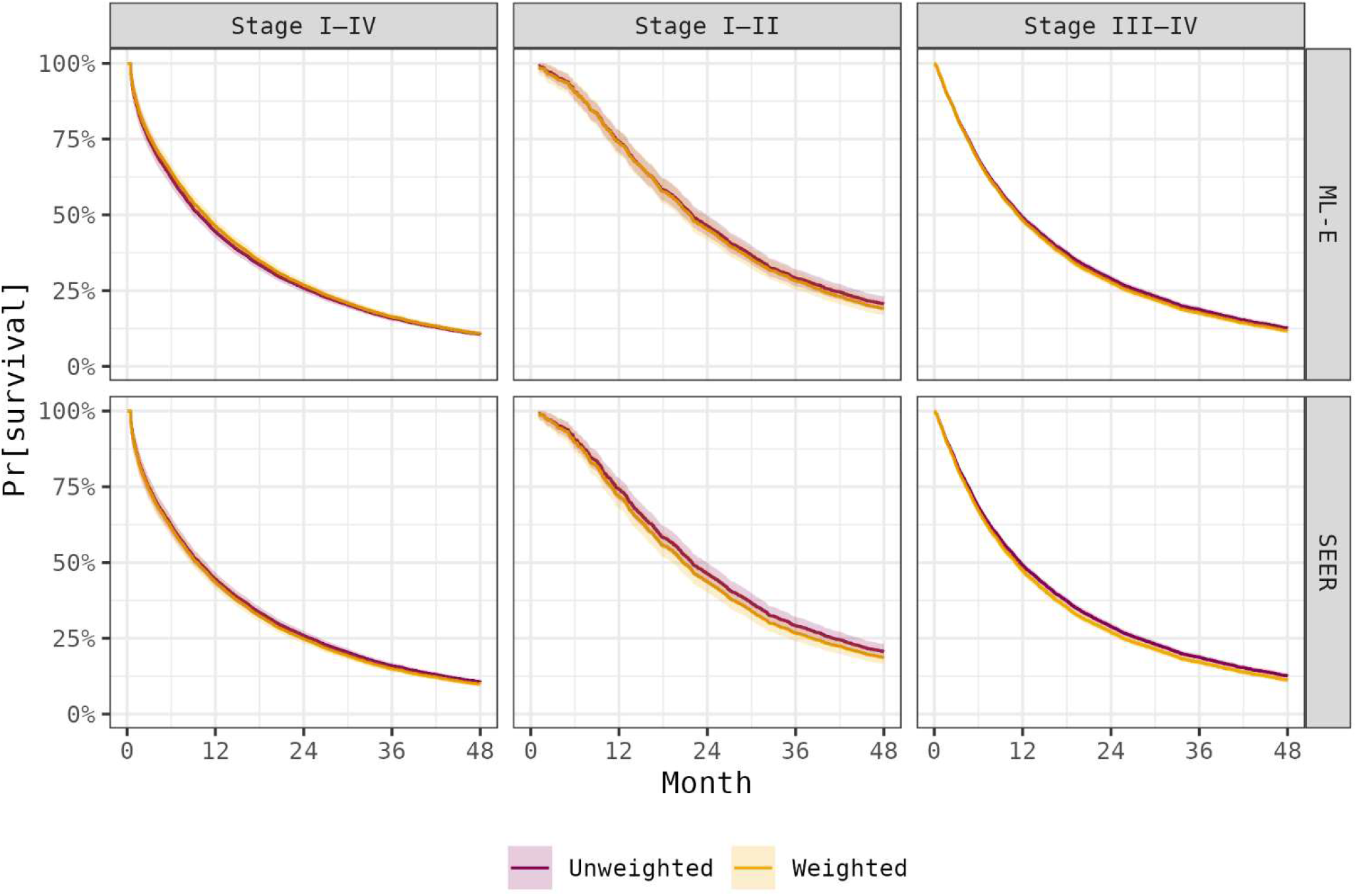
Marginal Real-world Overall Survival Estimated Within the Selected CGDB Sample And Extrapolated To The SEER Or ML-E Referent Populations With *W*^*S*^ Sampling. Time zero was the date of diagnosis for stages I–IV & I–II and the date of initiating first-line systemic therapy for the stages III–IV.

## DISCUSSION

Transportability analyses enables the use of combined information on the underlying target population to extrapolate to the unobserved counterfactual had the target population been under treatment. We show, by standardizing the joint distributions of effect modifiers within a selected sample to that of SEER cancer registrations, how the SATE for a USA Clinico-Genomic Database was equivalent to the PATE because the joint distributions were not discernibly different between the selected & unselected. We further demonstrate the absence of selection bias through a quantitative framework to assess the potential bias introduced due to non-random selection through each process that, unlike the ASD, has a direct influence on a weighted estimator.

The unified definition of selection bias is ‘any bias away from the true causal effect in the referent population, due to selecting the sample from the referent population.’^4^ Our vignette showed that the SATE was an unbiased estimation of the PATE, thereby suggesting the absence of any bias from selection through a function of requiring NGS and geographic sampling. However, unbiased sample estimates should not be assumed and transportability analyses can still be worthwhile for obtaining valid uncertainty bounds for the population. Potential issues to consider beforehand include having data for a valid target population — whether nested or non-nested for generalizability & transportability, respectively ^15^ — an effective sample size sufficient for precise confidence intervals ^21,22^, and the measurement of an harmonised set of effect modifiers required to achieve exchangeability under selection mutual to both sample and target populations ^9^.

We further demonstrate a quantitative framework to illustrate the potential bias introduced due to non-random selection by stepwise comparison of effect modifiers through each selection process. According to this framework, the non-random selection did not cause a differential selection of effect modifiers. In expectation, standardizing the differences in baseline characteristics between the selected and unselected had no influence on estimates of overall survival. This was despite the ASD suggesting imbalances in some marginal distributions. Naturally the *β* has a different yardstick for determining differences than the ASD. Nonetheless, comparing the marginal distributions has some limitations: knowing that the means of two — preferably Gaussian — marginal distributions are within 0.1 standard deviations does not inform on how a weighted estimator of the counterfactual may improve our inferences over a naïve estimator calculated within a convenience sample. Biases caused by non-random sample selection are proportional to a function of differing covariate distributions and strong effect modification. The Bhattacharyya coefficient is a quantitative measure of the former, assessing the closeness of two probability distributions; a higher *β* being indicative of a lesser reliance on extrapolation of the counterfactual. Future work should simulate heuristics for the interpretation of *β* under differing distributions and varying strengths of effect modification. Overall, our findings are consistent with past studies comparing Flatiron Health databases with cancer registrations ^23,24^ but its potential for bias has not hitherto been addressed.

Our study has some limitations. First, in the absence of population-wide cancer registrations, we assume that SEER registrations for participating states covering 34.6% of the USA population is a valid representation of the target population ^25^. Notably, SEER includes a higher proportion of the geriatric population less likely to receive treatment ^24^ and there is variability in the distributions of race across participating states ^26^. Secondly, transportability of ATEs and the *β* require no model mis-specification of effect modifiers in the respective models ^9,27,28^. Although exchangeability cannot be empirically tested, causal Directed Acyclic Graphs ^29^ and Quantitative Bias Analyses ^30,31^ are useful guiding tools. Finally, for simplicity we make several assumptions; namely that data was Missing Completely at Random — thereby justifying excluding missing data — and that delayed entry and loss to follow-up was random. If data were Missing At Random — poor prognosis leading to unnecessary diagnostic work-up in ascertaining stage, histological classification, for example — then this exclusion would skew study and referent population characteristics toward earlier disease by disproportionally excluding late-stage cancers. If there existed any violations to dependent left-truncation bias whereby progression of disease after diagnosis is causative of either ordering NGS ^6,18^ or informative censoring on last clinical activity after which death cannot be ascertained ^3,32^, further product weights should be derived from time-varying covariates measured post baseline ^14,33^.

In summary, combined information from cancer registrations can be used to extend inferences from a selected sample to the underlying target population. We show that estimates within a USA Clinico-Genomic Database were an unbiased estimate of the population because the sequential selection did not differentially select effect modifiers causative of survival.

## Data Availability

The data that support the findings of this study have been originated by Flatiron Health, Inc. Requests for data sharing by license or by permission for the specific purpose of replicating results in this manuscript can be submitted to. Surveillance, Epidemiology and End Results (SEER) data is publicly available with a signed data-use agreement [https://seer.cancer.gov/data]

---

**Supporting Table 1:** ICD-O-2 Codes Used For Histological Confirmation And Classification Of Non-Small Cell Lung Cancer.

| **Histological classification** | **ICD-O-2 codes** |
| --- | --- |
| Non-squamous cell carcinoma | **Defined as Adenocarcinomas, Large-cell carcinomas or Other specified carcinomas**<br>**Adenocarcinomas:**<br>8015, 8050, 8140, 8143, 8147, 8190, 8201, 8211, 8250, 8260, 8290, 8310, 8320, 8323, 8333, 8401, 8440, 8470, 8480, 8490, 8503, 8507, 8550, 8570, 8574, 8576<br>**Large-cell carcinomas:**<br>8012, 8021, 8034, 8082<br>**Other specified carcinomas:**<br>8003, 8022, 8030, 8035, 8200, 8240, 8243, 8249, 8430, 8525, 8560, 8562, 8575 |
| Squamous cell carcinoma | 8051, 8070, 8078, 8083, 8090, 8094, 8120, 8123 |

**Supporting Table 2:** Effect Modifiers for Stages I–IV CGDB, ML-E, And SEER Populations Before Exclusion Of Missing Data.

| **Effect Modifier** | **CGDB**<br>**(n 17,230)** | **ML-E**<br>**(n 199,278)** | **SEER**<br>**(n 240,943)** |
| --- | --- | --- | --- |
| Age at diagnosis |  |  |  |
| Median<br>(Interquartile range) | 68.0<br>(61.0, 75.0) | 70.0<br>(63.0, 76.0) | 70.0<br>(62.0, 77.0) |
| Unknown | 0<br>(0.0%) | 1<br>(0.0%) | 0<br>(0.0%) |
| Gender |  |  |  |
| Female | 8,793<br>(51.0%) | 98,928<br>(49.6%) | 114,973<br>(47.7%) |
| Male | 8,437<br>(49.0%) | 100,334<br>(50.3%) | 125,970<br>(52.3%) |
| Unknown | 0<br>(0.0%) | 16<br>(0.0%) | 0<br>(0.0%) |
| Race |  |  |  |
| Black | 1,140<br>(6.6%) | 15,898<br>(8.0%) | 28,263<br>(11.7%) |
| Other | 2,944<br>(17.1%) | 29,901<br>(15.0%) | 18,023<br>(7.5%) |
| White | 11,600<br>(67.3%) | 134,421<br>(67.5%) | 193,988<br>(80.5%) |
| Unknown | 1,546<br>(9.0%) | 19,058<br>(9.6%) | 669<br>(0.3%) |
| AJCC stage |  |  |  |
| Stage I | 1,849<br>(10.7%) | 44,729<br>(22.4%) | — |
| Stage II | 1,290<br>(7.5%) | 18,164<br>(9.1%) | — |
| Stage III | 3,938<br>(22.9%) | 43,747<br>(22.0%) | — |
| Stage IV | 9,518<br>(55.2%) | 80,883<br>(40.6%) | — |
| Unknown | 635<br>(3.7%) | 11,755<br>(5.9%) | — |
| SEER stage<sup>1</sup> |  |  |  |
| Localized | 3,139<br>(18.2%) | 62,893<br>(31.6%) | 55,754<br>(23.1%) |
| Regional | 3,938<br>(22.9%) | 43,747<br>(22.0%) | 56,664<br>(23.5%) |
| Distant | 9,518<br>(55.2%) | 80,883<br>(40.6%) | 122,410<br>(50.8%) |
| Unknown | 635<br>(3.7%) | 11,755<br>(5.9%) | 6,115<br>(2.5%) |
| Histological classification |  |  |  |
| Non-squamous cell carcinoma | 12,814<br>(74.4%) | 131,339<br>(65.9%) | 144,502<br>(60.0%) |
| Squamous cell carcinoma | 3,727<br>(21.6%) | 58,273<br>(29.2%) | 58,637<br>(24.3%) |
| Unknown | 689<br>(4.0%) | 9,666<br>(4.9%) | 37,804<br>(15.7%) |

Abbreviations: CGDB, Clinico-Genomic Database [population]; ML-E, Machine Learning-Extracted [population]; SEER, Surveillance, Epidemiology, and End Results [population]; AJCC, American Joint Committee on Cancer
<sup>1</sup> SEER stage approximated for CGDB & ML-E populations

**Supporting Table 3:**
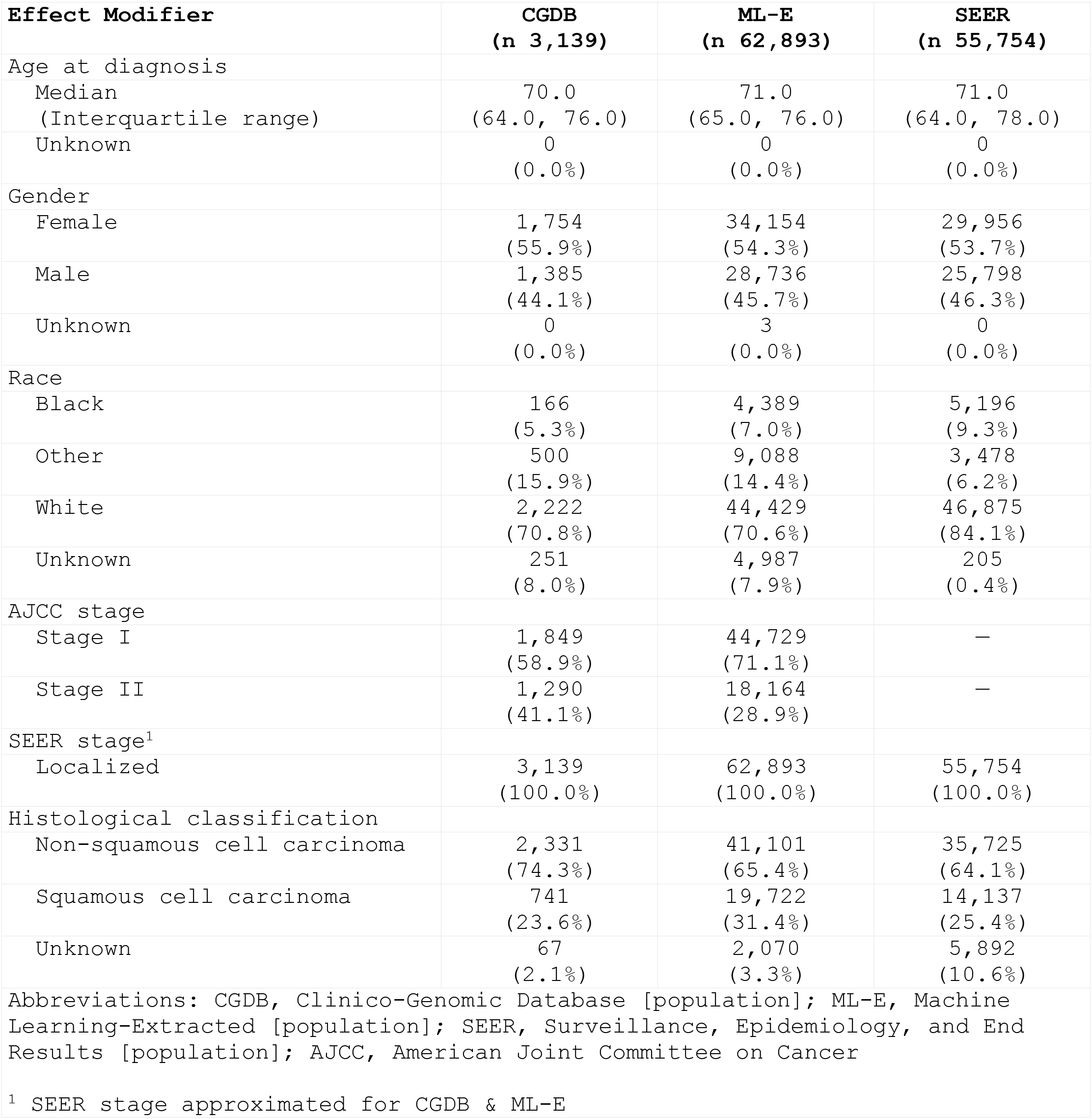
Effect Modifiers For Stages I–II CGDB, ML-E, And SEER Populations Before Exclusion Of Missing Data.

| **Effect Modifier** | **CGDB**<br>**(n 3,139)** | **ML-E**<br>**(n 62,893)** | **SEER**<br>**(n 55,754)** |
| --- | --- | --- | --- |
| Age at diagnosis |  |  |  |
| Median<br>(Interquartile range) | 70.0<br>(64.0, 76.0) | 71.0<br>(65.0, 76.0) | 71.0<br>(64.0, 78.0) |
| Unknown | 0<br>(0.0%) | 0<br>(0.0%) | 0<br>(0.0%) |
| Gender |  |  |  |
| Female | 1,754<br>(55.9%) | 34,154<br>(54.3%) | 29,956<br>(53.7%) |
| Male | 1,385<br>(44.1%) | 28,736<br>(45.7%) | 25,798<br>(46.3%) |
| Unknown | 0<br>(0.0%) | 3<br>(0.0%) | 0<br>(0.0%) |
| Race |  |  |  |
| Black | 166<br>(5.3%) | 4,389<br>(7.0%) | 5,196<br>(9.3%) |
| Other | 500<br>(15.9%) | 9,088<br>(14.4%) | 3,478<br>(6.2%) |
| White | 2,222<br>(70.8%) | 44,429<br>(70.6%) | 46,875<br>(84.1%) |
| Unknown | 251<br>(8.0%) | 4,987<br>(7.9%) | 205<br>(0.4%) |
| AJCC stage |  |  |  |
| Stage I | 1,849<br>(58.9%) | 44,729<br>(71.1%) | — |
| Stage II | 1,290<br>(41.1%) | 18,164<br>(28.9%) | — |
| SEER stage<sup>1</sup> |  |  |  |
| Localized | 3,139<br>(100.0%) | 62,893<br>(100.0%) | 55,754<br>(100.0%) |
| Histological classification |  |  |  |
| Non-squamous cell carcinoma | 2,331<br>(74.3%) | 41,101<br>(65.4%) | 35,725<br>(64.1%) |
| Squamous cell carcinoma | 741<br>(23.6%) | 19,722<br>(31.4%) | 14,137<br>(25.4%) |
| Unknown | 67<br>(2.1%) | 2,070<br>(3.3%) | 5,892<br>(10.6%) |

Abbreviations: CGDB, Clinico-Genomic Database [population]; ML-E, Machine Learning-Extracted [population]; SEER, Surveillance, Epidemiology, and End Results [population]; AJCC, American Joint Committee on Cancer
<sup>1</sup> SEER stage approximated for CGDB & ML-E

**Supporting Table 4:** Effect Modifiers For Stages III–IV CGDB, ML-E, And SEER Populations Before Exclusion Of Missing Data.

| **Effect Modifier** | **CGDB**<br>**(n 13,456)<sup>1</sup>** | **ML-E**<br>**(n 124,630)** | **SEER**<br>**(n 179,074)** |
| --- | --- | --- | --- |
| Age at diagnosis |  |  |  |
| Median (Interquartile range) | 68.0<br>(60.0, 75.0) | 69.0<br>(61.0, 75.0) | 69.0<br>(61.0, 77.0) |
| Unknown | 0<br>(0.0%) | 0<br>(0.0%) | 0<br>(0.0%) |
| Gender |  |  |  |
| Female | 6,709<br>(49.9%) | 58,969<br>(47.3%) | 82,093<br>(45.8%) |
| Male | 6,747<br>(50.1%) | 65,650<br>(52.7%) | 96,981<br>(54.2%) |
| Unknown | 0<br>(0.0%) | 11<br>(0.0%) | 0<br>(0.0%) |
| Race |  |  |  |
| Black | 931<br>(6.9%) | 10,484<br>(8.4%) | 22,415<br>(12.5%) |
| Other | 2,315<br>(17.2%) | 19,022<br>(15.3%) | 13,953<br>(7.8%) |
| White | 8,986<br>(66.8%) | 82,601<br>(66.3%) | 142,347<br>(79.5%) |
| Unknown | 1,224<br>(9.1%) | 12,523<br>(10.0%) | 359<br>(0.2%) |
| AJCC stage |  |  |  |
| Stage III | 3,938<br>(29.3%) | 43,747<br>(35.1%) | — |
| Stage IV | 9,518<br>(70.7%) | 80,883<br>(64.9%) | — |
| SEER stage<sup>2</sup> |  |  |  |
| Regional | 3,938<br>(29.3%) | 43,747<br>(35.1%) | 56,664<br>(31.6%) |
| Distant | 9,518<br>(70.7%) | 80,883<br>(64.9%) | 122,410<br>(68.4%) |
| Histological classification |  |  |  |
| Non-squamous cell carcinoma | 10,097<br>(75.0%) | 84,169<br>(67.5%) | 106,210<br>(59.3%) |
| Squamous cell carcinoma | 2,783<br>(20.7%) | 34,138<br>(27.4%) | 43,147<br>(24.1%) |
| Unknown | 576<br>(4.3%) | 6,323<br>(5.1%) | 29,717<br>(16.6%) |

Abbreviations: CGDB, Clinico-Genomic Database [population]; ML-E, Machine Learning-Extracted [population]; SEER, Surveillance, Epidemiology, and End Results [population]; AJCC, American Joint Committee on Cancer
<sup>1</sup> Stage III-IV patients that also initiated systemic therapy
<sup>2</sup> SEER stage approximated for CGDB & ML-E

**Supporting Table 5:** Effect Modifiers For Stages I–II ML-E & SEER Populations.

| **Effect Modifier** | **ML-E**<br>**(n 56,031)** | **SEER**<br>**(n 49,685)** | **ASD** |
| --- | --- | --- | --- |
| Age at diagnosis<sup>1</sup> | 71.0<br>(65.0,<br>76.0) | 71.0<br>(64.0,<br>78.0) | 0.07 |
| Gender |  |  | 0.00 |
| Female | 30,453<br>(54.4%) | 26,894<br>(54.1%) |  |
| Male | 25,578<br>(45.6%) | 22,791<br>(45.9%) |  |
| Race |  |  | 0.30 |
| Black | 4,250<br>(7.6%) | 4,637<br>(9.3%) |  |
| Other | 8,827<br>(15.8%) | 3,185<br>(6.4%) |  |
| White | 42,954<br>(76.7%) | 41,863<br>(84.3%) |  |
| SEER stage |  |  | 0.00 |
| Localized | 56,031<br>(100.0%) | 49,685<br>(100.0%) |  |
| Histological classification |  |  | 0.09 |
| Non-squamous cell carcinoma | 37,856<br>(67.6%) | 35,578<br>(71.6%) |  |
| Squamous cell carcinoma | 18,175<br>(32.4%) | 14,107<br>(28.4%) |  |
| Abbreviations: ML-E, Machine Learning-Extracted [population]; SEER, Surveillance, Epidemiology, and End Results [population]; ASD, Absolute Standardized Difference |  |  |  |
| <sup>1</sup> Median (Interquartile range) |  |  |  |

**Supporting Table 6:**
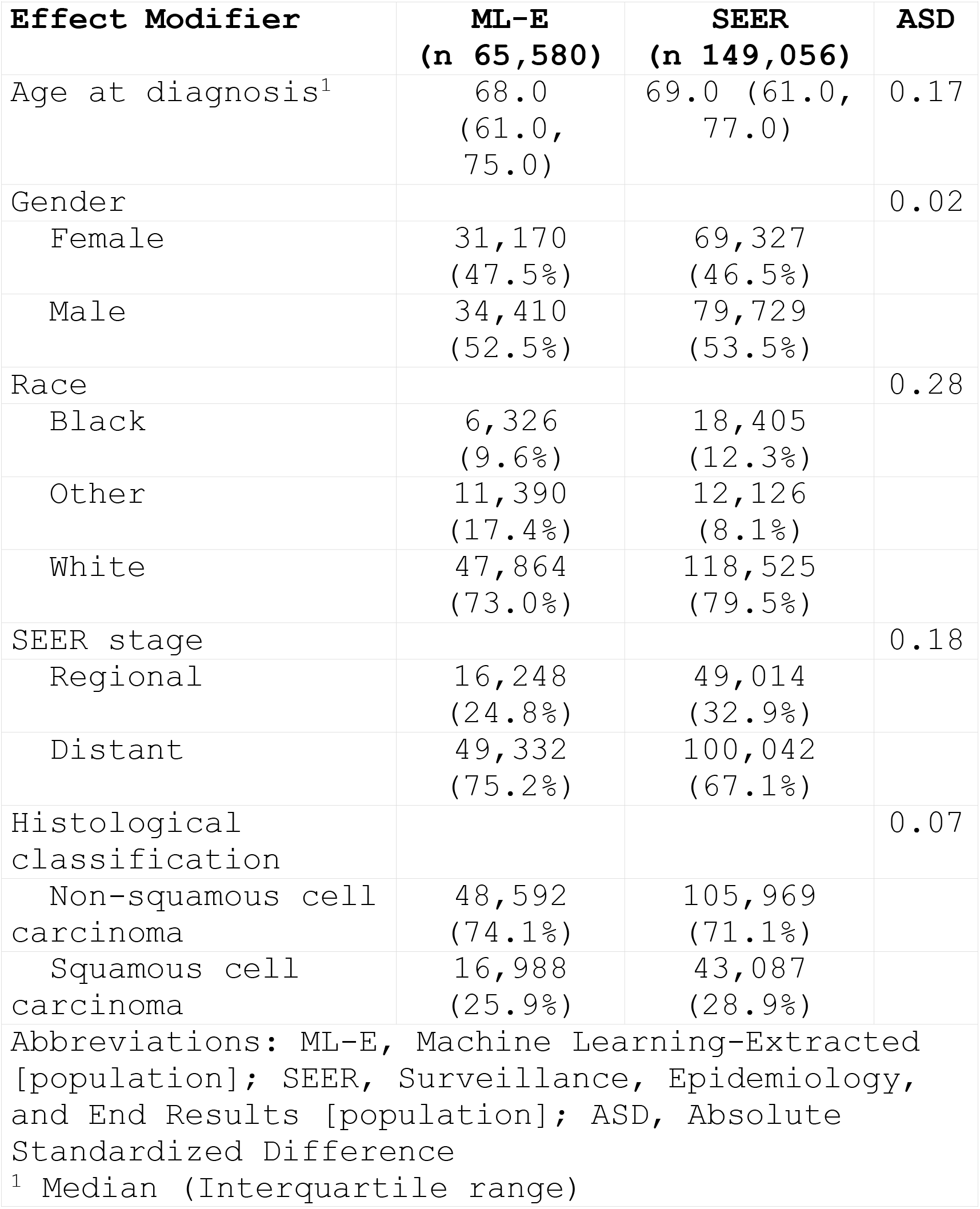
Effect Modifiers For Stages III–IV ML-E & SEER Populations.

**Supporting Table 7:** Effect Modifiers For Stages I–II CGDB & ML-E Populations.

| Effect Modifier | Unweighted |  |  | *W<sup>S</sup>*-Weighted |  |  |
| --- | --- | --- | --- | --- | --- | --- |
|  | CGDB<br>(n 2,826) | ML-E<br>(n 56,031) | ASD | CGDB<br>(n 2,833)<sup>1</sup> | ML-E<br>(n 56,031) | ASD |
| Age at diagnosis<sup>1</sup> | 70.0<br>(64.0,<br>76.0) | 71.0<br>(65.0,<br>76.0) | 0.09 | 71.0<br>(65.0,<br>76.0) | 71.0<br>(65.0,<br>76.0) | 0.01 |
| Gender |  |  | 0.03 |  |  | 0.02 |
| Female | 1,583<br>(56.0%) | 30,453<br>(54.4%) |  | 1,515<br>(53.5%) | 30,453<br>(54.4%) |  |
| Male | 1,243<br>(44.0%) | 25,578<br>(45.6%) |  | 1,317<br>(46.5%) | 25,578<br>(45.6%) |  |
| Race |  |  | 0.08 |  |  | 0.01 |
| Black | 165<br>(5.8%) | 4,250<br>(7.6%) |  | 219<br>(7.7%) | 4,250<br>(7.6%) |  |
| Other | 489<br>(17.3%) | 8,827<br>(15.8%) |  | 453<br>(16.0%) | 8,827<br>(15.8%) |  |
| White | 2,172<br>(76.9%) | 42,954<br>(76.7%) |  | 2,161<br>(76.3%) | 42,954<br>(76.7%) |  |
| AJCC stage |  |  | 0.26 |  |  | 0.00 |
| Stage I | 1,658<br>(58.7%) | 39,761<br>(71.0%) |  | 2,015<br>(71.2%) | 39,761<br>(71.0%) |  |
| Stage II | 1,168<br>(41.3%) | 16,270<br>(29.0%) |  | 817<br>(28.8%) | 16,270<br>(29.0%) |  |
| Histological classification |  |  | 0.18 |  |  | 0.02 |
| Non-squamous cell carcinoma | 2,143<br>(75.8%) | 37,856<br>(67.6%) |  | 1,889<br>(66.7%) | 37,856<br>(67.6%) |  |
| Squamous cell carcinoma | 683<br>(24.2%) | 18,175<br>(32.4%) |  | 944<br>(33.3%) | 18,175<br>(32.4%) |  |

Abbreviations: CGDB, Clinico-Genomic Database [population]; ML-E, Machine Learning-Extracted [population]; ASD, Absolute Standardized Difference; AJCC, American Joint Committee on Cancer
<sup>1</sup> Effective Sample Size of 2,485
<sup>2</sup> Median (Interquartile range)

**Supporting Table 8:** Effect Modifiers For Stages III–IV CGDB & ML-E Populations.

| Effect Modifier | Unweighted |  |  | *W<sup>S</sup>*-Weighted |  |  |
| --- | --- | --- | --- | --- | --- | --- |
|  | CGDB<br>(n 9,172)<sup>1</sup> | ML-E<br>(n 106,546) | ASD | CGDB<br>(n 9,163)<sup>2</sup> | ML-E<br>(n 106,546) | ASD |
| Age at diagnosis<sup>3</sup> | 67.0<br>(60.0,<br>74.0) | 69.0<br>(61.0,<br>75.0) | 0.13 | 68.0<br>(61.0,<br>75.0) | 69.0<br>(61.0,<br>75.0) | 0.00 |
| Gender |  |  | 0.06 |  |  | 0.01 |
| Female | 4,618<br>(50.3%) | 50,653<br>(47.5%) |  | 4,318<br>(47.1%) | 50,653<br>(47.5%) |  |
| Male | 4,554<br>(49.7%) | 55,893<br>(52.5%) |  | 4,845<br>(52.9%) | 55,893<br>(52.5%) |  |
| Race |  |  | 0.08 |  |  | 0.01 |
| Black | 686<br>(7.5%) | 9,938<br>(9.3%) |  | 863<br>(9.4%) | 9,938<br>(9.3%) |  |
| Other | 1,776<br>(19.4%) | 18,188<br>(17.1%) |  | 1,579<br>(17.2%) | 18,188<br>(17.1%) |  |
| White | 6,710<br>(73.2%) | 78,420<br>(73.6%) |  | 6,721<br>(73.4%) | 78,420<br>(73.6%) |  |
| AJCC stage<sup>1</sup> |  |  | 0.24 |  |  | 0.01 |
| Stage III | 2,278<br>(24.8%) | 38,079<br>(35.7%) |  | 3,238<br>(35.3%) | 38,079<br>(35.7%) |  |
| Stage IV | 6,894<br>(75.2%) | 68,467<br>(64.3%) |  | 5,924<br>(64.7%) | 68,467<br>(64.3%) |  |
| Histological classification |  |  | 0.20 |  |  | 0.01 |
| Non-squamous cell carcinoma | 7,297<br>(79.6%) | 75,786<br>(71.1%) |  | 6,544<br>(71.4%) | 75,786<br>(71.1%) |  |
| Squamous cell carcinoma | 1,875<br>(20.4%) | 30,760<br>(28.9%) |  | 2,619<br>(28.6%) | 30,760<br>(28.9%) |  |

Abbreviations: CGDB, Clinico-Genomic Database [population]; ML-E, Machine Learning-Extracted [population]; ASD, Absolute Standardized Difference; AJCC, American Joint Committee on Cancer
<sup>1</sup> Stage III-IV patients that also initiated systemic therapy
<sup>2</sup> Effective Sample Size of 8,266
<sup>3</sup> Median (Interquartile range)

**Supporting Table 9:**
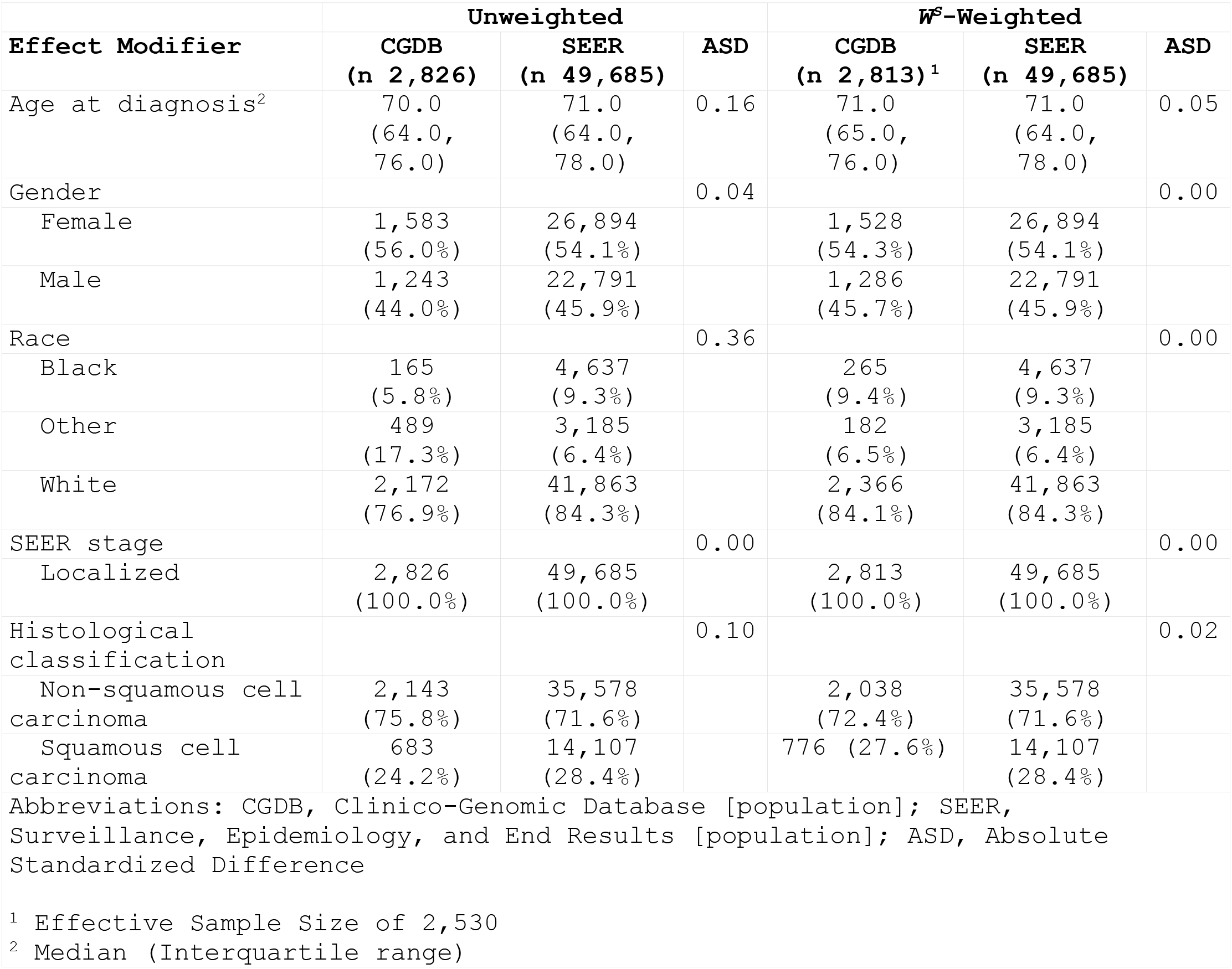
Effect Modifiers For Stages I–II CGDB & SEER Populations.

**Supporting Table 10:** Effect Modifiers For Stages III–IV CGDB & SEER Populations.

| Effect Modifier | Unweighted |  |  | W<sup>s</sup>-Weighted |  |  |
| --- | --- | --- | --- | --- | --- | --- |
|  | CGDB<br>(n 9,172) | SEER<br>(n 149,056) | ASD | CGDB<br>(n 9,088)<sup>1</sup> | SEER<br>(n 149,056) | ASD |
| Age at diagnosis<sup>2</sup> | 67.0<br>(60.0,<br>74.0) | 69.0<br>(61.0,<br>77.0) | 0.24 | 69.0<br>(62.0,<br>76.0) | 69.0<br>(61.0,<br>77.0) | 0.06 |
| Gender |  |  | 0.08 |  |  | 0.01 |
| Female | 4,618<br>(50.3%) | 69,327<br>(46.5%) |  | 4,172<br>(45.9%) | 69,327<br>(46.5%) |  |
| Male | 4,554<br>(49.7%) | 79,729<br>(53.5%) |  | 4,916<br>(54.1%) | 79,729<br>(53.5%) |  |
| Race |  |  | 0.35 |  |  | 0.02 |
| Black | 686<br>(7.5%) | 18,405<br>(12.3%) |  | 1,145<br>(12.6%) | 18,405<br>(12.3%) |  |
| Other | 1,776<br>(19.4%) | 12,126<br>(8.1%) |  | 776<br>(8.5%) | 12,126<br>(8.1%) |  |
| White | 6,710<br>(73.2%) | 118,525<br>(79.5%) |  | 7,167<br>(78.9%) | 118,525<br>(79.5%) |  |
| SEER stage |  |  | 0.18 |  |  | 0.01 |
| Regional | 2,278<br>(24.8%) | 49,014<br>(32.9%) |  | 2,956<br>(32.5%) | 49,014<br>(32.9%) |  |
| Distant | 6,894<br>(75.2%) | 100,042<br>(67.1%) |  | 6,131<br>(67.5%) | 100,042<br>(67.1%) |  |
| Histological classification |  |  | 0.20 |  |  | 0.01 |
| Non-squamous cell carcinoma | 7,297<br>(79.6%) | 105,969<br>(71.1%) |  | 6,516<br>(71.7%) | 105,969<br>(71.1%) |  |
| Squamous cell carcinoma | 1,875<br>(20.4%) | 43,087<br>(28.9%) |  | 2,571<br>(28.3%) | 43,087<br>(28.9%) |  |

Abbreviations: CGDB, Clinico-Genomic Database [population]; SEER, Surveillance, Epidemiology, and End Results [population]; ASD, Absolute Standardized Difference
<sup>1</sup> Effective Sample Size of 7,601
<sup>2</sup> Median (Interquartile range)

**Supporting Table 11:** Sensitivity Analysis Comparing Effect Modifiers for Stages I–IV CGDB & SEER Populations Harmonized To The Same Study Period (2011–2016)

| Effect Modifier | Unweighted |  | ASD<sup>2</sup> |
| --- | --- | --- | --- |
|  | CGDB<br>(n 5,249) | SEER<br>(n 198,741) |  |
| Age at diagnosis<sup>1</sup> | 66.0<br>(58.0, 73.0) | 70.0<br>(62.0, 77.0) | 0.41 |
| Gender |  |  | 0.10 |
| Female | 2,813<br>(53.6%) | 96,221<br>(48.4%) |  |
| Male | 2,436<br>(46.4%) | 102,520<br>(51.6%) |  |
| Race |  |  | 0.33 |
| Black | 325<br>(6.2%) | 23,042<br>(11.6%) |  |
| Other | 902<br>(17.2%) | 15,311<br>(7.7%) |  |
| White | 4,022<br>(76.6%) | 160,388<br>(80.7%) |  |
| SEER stage |  |  | 0.02 |
| Localized | 1,353<br>(25.8%) | 49,685<br>(25.0%) |  |
| Regional | 1,293<br>(24.6%) | 49,014<br>(24.7%) |  |
| Distant | 2,603<br>(49.6%) | 100,042<br>(50.3%) |  |
| Histological classification |  |  | 0.24 |
| Non-squamous cell carcinoma | 4,279<br>(81.5%) | 141,547<br>(71.2%) |  |
| Squamous cell carcinoma | 970<br>(18.5%) | 57,194<br>(28.8%) |  |

Abbreviations: CGDB, Clinico-Genomic Database [population]; SEER, Surveillance, Epidemiology, and End Results [population]; ASD, Absolute Standardized Difference
<sup>1</sup> Median (Interquartile range)
<sup>2</sup> $\beta$ was 0.95

## Supporting File 1: Causal Assumptions

We present our causal assumptions in Supplementary Figure 1, where *A* represents a binary treatment, *Y* the outcome of Real-world Overall Survival, *Z* a set of baseline variables (age, gender, race, stage, histological classification), *T* a binary node indicating having undergone Next Generation Sequencing (NGS), and *S* a node indicating selection into the sample [from the target population]. A [box] depicts conditioning through experimental design (restriction where *S = 1*). The causal and non-causal paths are depicted as dashed & red lines, respectively.

**Supporting Figure 1:**
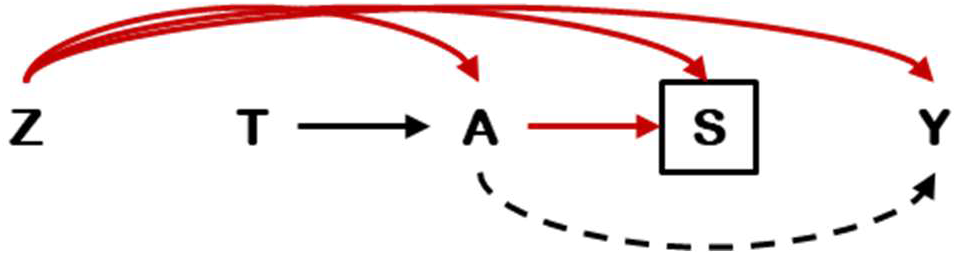
Directed Acyclic Graph showing the causal assumptions underlying the study of a convenience CGDB sample.

Selection into the sample from the SEER target population was assumed *a priori* to be a function of geographic sampling and further eligibility criteria. Note that while in real-world clinical practice a set of baseline variables *Z* can cause the ordering of NGS *T*, in our example *T* is defined only by the population. SEER patients cannot by definition undergo NGS. Nevertheless, any differential selection of patient subgroups through the requirement of NGS is corrected downstream by weighting on the Inverse Odds of Selection. Separately, undergoing NGS *T* only affects the outcome *Y* through directing treatment *A* based on the tumor genotype.

In the language of causal Directed Acyclic Graphs ^1^, a type-1 selection bias (collider-restriction bias) results from conditioning on a common effect of both the treatment (or cause of treatment) and the outcome (or cause of the outcome) ^2,3^. Conditioning on a collider variable by studying a restricted cohort can induce a spurious association between its parents (treatment and outcome) even if they are marginally independent. This collider bias can inflate, attenuate or even reverse-sign associations ^4,5^. Where type-1 selection biases is a result of conditioning on a common effect of *A & Y* (*A* > [*Z*] < *Y*), confounding biases are defined by open backdoor paths dealing with common causes of *A & Y* (*A* < *Z* > *Y*).

The graph above illustrates three paths:

— a causal path (*A* > *Y*) which we wish to isolate;
— a non-causal confounding path (*A* < *Z* > *Y*);
— a non-causal collider-restriction path (*A* > [*S*] < *Z* > *Y*), invoked by restricting on a collider variable by restricting on levels of *S* — a common effect of *Z* and treatment *A* (i.e. type-1 selection bias)

### Non-causal confounding path

Sampling with Inverse/Stabilized Probability of Treatment Weights (*W*^*A*^) creates a pseudo-population in which the treatment received is conditionally independent of the outcome (under the null), through weighting on the conditional probability of treatment received given a set of covariates *Z* that satisfy the backdoor criteria (*P[A = a*|*Z = z]*). In effect, there is no arrow from *Z* to *A* in this *W*^*A*^-weighted population.

### Non-causal collider-restriction path

Sampling with Transportability Weights (*W*^*S*^) creates a pseudo-population in which selection is conditionally independent of the outcome, through weighting on the conditional probability or odds of selection given a set of covariates *Z* that cause selection. In effect, there is no arrow from *S* to *Y* in this *W*^*S*^-weighted population. Under a nested sampling structure — under which the sampling fraction is knowable since we all selected samples are identifiable within the referent population extending — extending inferences becomes an issue of generalizability requiring probability weights ^6^. Under a non-nested sampling design, the sampling fraction is unknowable since we are missing information on the entire unselected population (where *S=0*), extending inferences becomes an issue of transportability requiring odds weights. The sampling fraction is also unknowable under a full nested design if the selected sample is not directly identifiable within the referent population — as is the case that for data protection reasons Flatiron Health Clinico-Genomic Database (CGDB) patients are not identifiable within the Flatiron health Machine learning Extracted (ML-E) database.

### Average Treatment Effects

In the estimation of the Sample Average Treatment Effect (SATE), only the confounding path is blocked with a *W*^*A*^-weighted estimator, leaving open the collider-restriction path as shown in Supporting Figure 2. When we wish to extend the SATE to the PATE, we the product terms of treatment & selection weights (*W*^*A*^*W*^*S*^) as shown in Supporting Figure 3.

**Supporting Figure 2:**
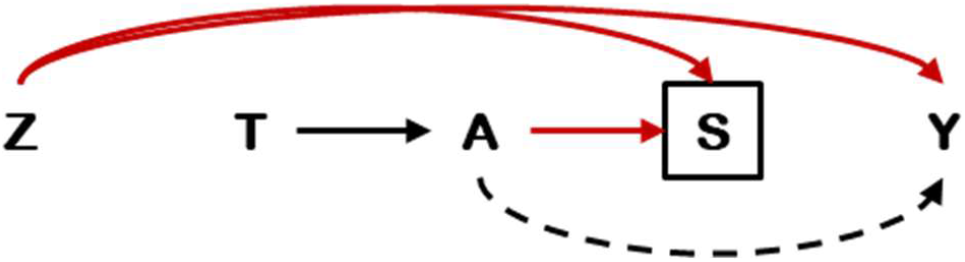
Identification of the Sample Average Treatment Effect (SATE). Note the presence of a non-causal collider-restriction path.

**Supporting Figure 3:**
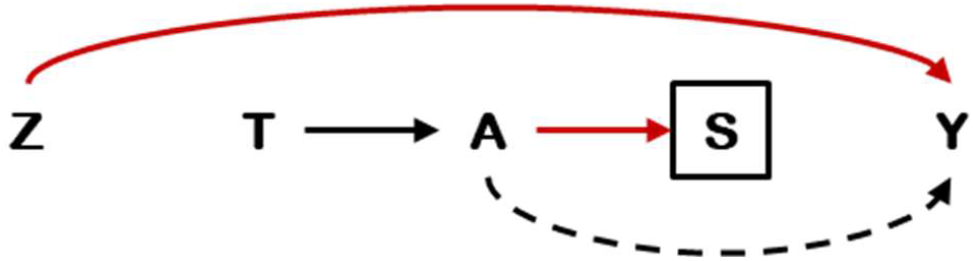
Identification of the Population Average Treatment Effect (PATE). Note the closure of non-causal paths.

## Acknowledgements

The authors thank Miguel Miranda (AstraZeneca) for his involvement in discussions on statistical analyses

